# *In Silico* Network Perturbation Reveals Hierarchical Roles of DNA Repair and Glycosylation Linking Exercise to Human Ageing Clocks

**DOI:** 10.64898/2026.02.01.26345311

**Authors:** Ciara G. Juan, Lazaros Ntasis

**Affiliations:** Faculty of Life and Health Sciences, Ulster University, Belfast, BT15 1ED; Department of Sport Organisation and Management, University of Peloponnese, Sparta, Greece 231 00

**Keywords:** In silico perturbation, exercise, ageing, ageing clocks, biological age, DNA repair, glycosylation, human study

## Abstract

Regular physical activity delays biological ageing, yet how transient exercise-induced molecular responses are translated into stable ageing signatures in humans remains unclear. Here, we introduce the first *in silico* perturbation framework for human exercise biology that integrates genetically anchored gene prioritisation, graph-based network modelling, and human experimental validation. Genes causally associated with habitual vigorous physical activity (VPA) were prioritised using Mendelian randomisation (MR) across proteomic, epigenomic, glycomic, and single-cell transcriptomic layers and represented using self-supervised graph learning. Targeted acute *in silico* perturbations were then propagated via network diffusion within a network shaped by genetically proxied habitual VPA to forecast downstream alignment with epigenetic and proteomic ageing clocks. Perturbation of validated glycosylation enzymes consistently yielded diffusion neighbourhoods enriched for clock-associated genes (empirical p < 0.01), whereas perturbation of canonical DNA repair and stress response genes did not consistently align with ageing clock architecture. Acute high-intensity exercise validation demonstrated rapid modulation of plasma glycosylation alongside activation of DNA repair programmes, providing biological context for distinct network behaviours. Together, these findings reveal a hierarchical organisation in which DNA repair pathways act as adaptive buffers of acute physiological stress but do not directly encode biological ageing state, while downstream glycosylation networks occupy a more proximal, integrative position, predictively encoding stable molecular states captured by human ageing clocks. By resolving stress buffering from ageing state encoding at the network level, this work refines damage-centric models of ageing and establishes *in silico* perturbation as a principled approach to forecast how habitual physical activity shapes long-term biological ageing trajectories.

Graphical Abstract
In silico perturbation framework linking habitual physical activity to ageing-related molecular architecture.
Genetically proxied habitual vigorous physical activity (VPA) is integrated with multi-omic Mendelian randomisation (MR) across epigenomic, transcriptomic, proteomic, and glycomic layers to construct a causal, exercise-adapted molecular network. Within this network, targeted acute in silico gene perturbations are propagated by network diffusion and evaluated for alignment with epigenetic and proteomic ageing clocks as external annotations. A controlled human high-intensity exercise intervention provides experimental validation, demonstrating acute activation of DNA repair programmes alongside rapid modulation of plasma glycosylation.

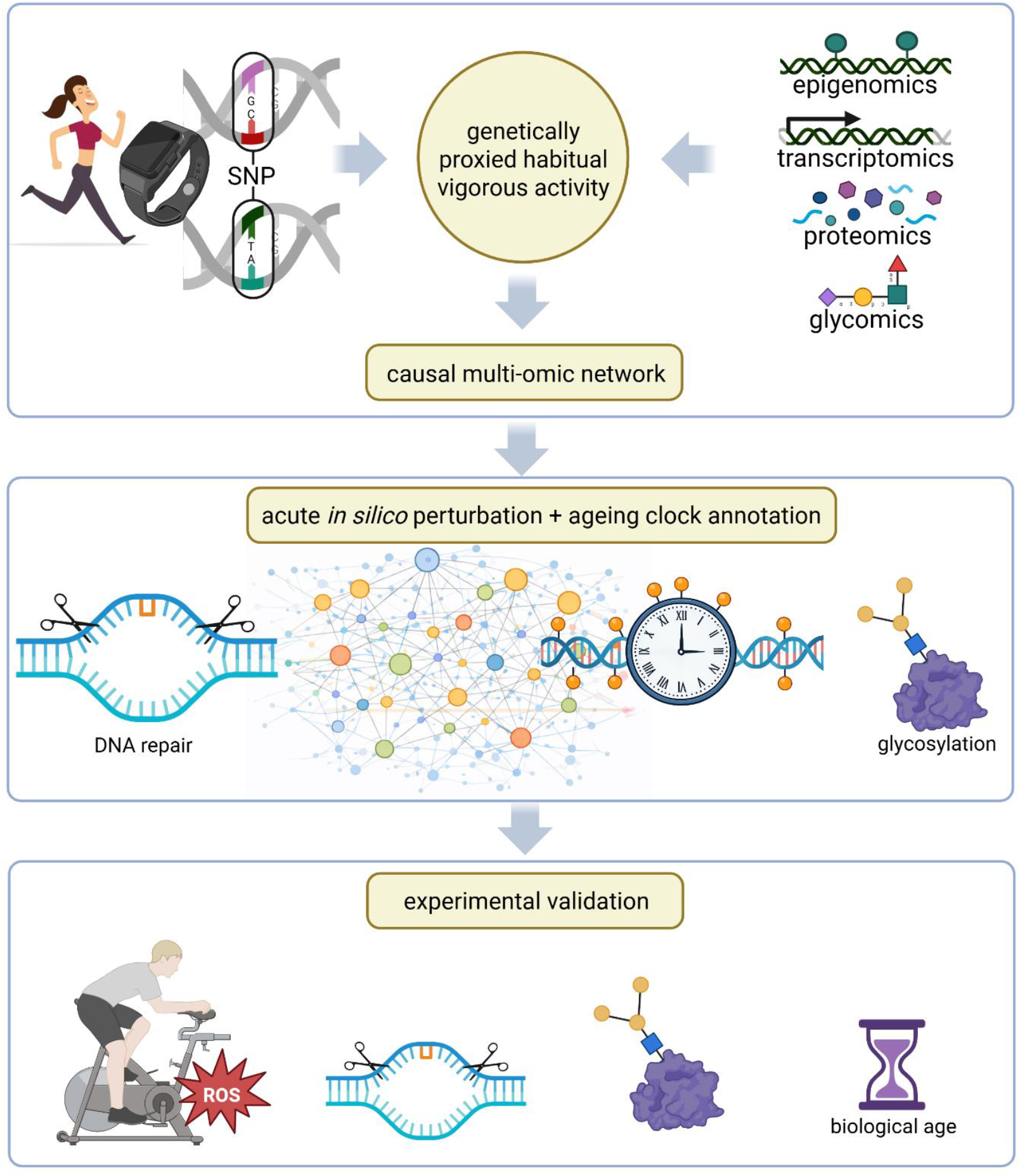

## Introduction

Regular physical activity is one of the most robust non-pharmacological interventions associated with delayed biological ageing and reduced risk of age-related disease. At the molecular level, exercise engages conserved stress-response programmes—including redox signalling, DNA damage sensing, and metabolic reprogramming—that promote cellular resilience and preserve genome integrity. In parallel, biological ageing clocks derived from epigenetic, proteomic, and glycomic data capture stable system-level molecular states that track chronological age and predict morbidity and mortality. Despite strong evidence linking exercise to both acute molecular responses and long-term ageing trajectories, the organisational hierarchy connecting transient exercise-induced stress responses to durable molecular ageing architecture in humans remains poorly defined.

DNA repair pathways occupy a central position in the immediate cellular response to exercise-induced physiological stress. Vigorous physical activity (VPA) transiently elevates reactive oxygen species (ROS) through increased mitochondrial flux, mechanical strain, and immune activation. These short-lived redox signals act as physiological cues that engage DNA damage sensing and repair pathways, including Poly(ADP-ribose) polymerase (PARP) and sirtuin-dependent mechanisms that coordinate genome maintenance with metabolic adaptation. Such responses are rapid, lesion-specific, and reversible, enabling repeated exposure to physiological stress without cumulative genomic instability (Radak et al., 2011; Williamson et al., 2020).

Protein glycosylation represents a fundamentally different layer of molecular regulation. Glycosylation is a ubiquitous post-translational modification that governs protein folding, stability, trafficking, immune recognition, and inflammatory signalling. Unlike acute stress-response pathways, glycan structures—particularly N-glycans on circulating plasma proteins and immunoglobulin G (IgG)—encode integrative information reflecting chronic immune activation, metabolic state, and inflammatory tone. Extensive glycomic evidence demonstrates that specific IgG N-glycan features, including galactosylation, sialylation, fucosylation, and bisecting GlcNAc, are tightly linked to inflammaging, cardiovascular disease, autoimmunity, and neurodegeneration, and shift predictably with biological ageing (Onigbinde et al., 2025). Mechanistically, glycosylation integrates upstream metabolic, inflammatory, and redox inputs via ROS-sensitive regulation of glycosyltransferase activity, Golgi and endoplasmic reticulum redox organisation, nucleotide-sugar availability and transport, and cytokine-driven signalling pathways, enabling dynamic but persistent remodelling of glycan structures in response to physiological stress (Kellokumpu, 2019; Khoder-Agha & Kietzmann, 2021). This positions glycosylation as a plausible molecular interface through which repeated physiological stress—such as habitual exercise—may be translated into stable ageing-related molecular states, rather than acting as an acute stress sensor itself.

Genetic approaches provide an entry point into resolving this hierarchy. Mendelian randomisation (MR) enables causal inference between habitual physical activity and molecular traits, allowing prioritisation of exercise-associated genes beyond observational correlation. However, most MR studies of exercise have focused on isolated genes or traits and have not resolved how exercise-responsive molecular features are organised within interacting networks. We recently addressed this limitation by integrating MR with supervised graph neural network to perform the first network-level prioritisation of exercise-associated genes across multiple molecular layers in humans (Juan & Ntasis, 2026). While this framework provided causal and mechanistic ranking, it remained inherently descriptive, identifying which genes are associated with habitual exercise without testing how perturbations propagate through molecular networks or whether such genes organise ageing-related molecular architecture.

To move beyond descriptive prioritisation, we apply network-based *in silico* perturbation within a molecular interaction network shaped by genetically proxied habitual VPA. Genes prioritised by the graph neural network are used in self-supervised graph representation learning, followed by MR-informed masked multi-task embedding refinement across proteomic, epigenomic, glycomic, and single-cell transcriptomic effect sizes. Single-event gene-level perturbations are then propagated to probe how acute molecular interventions reorganise downstream network architecture. Perturbation effects are quantified by measuring redistribution of network diffusion toward epigenetic ageing clock regions and proteomic ageing axes, providing a direct test of whether exercise-responsive pathways are positioned to shape stable ageing-related molecular states. Together, this framework integrates causal inference, self-supervised graph representation learning with multi-task MR-based embedding refinement, and acute *in silico* perturbation to distinguish transient stress-buffering programmes from downstream integrative pathways that encode long-term biological ageing in humans. Importantly, plasma and IgG glycosylation represent one of the most scalable and cost-effective omic layers for human validation, enabling direct experimental grounding of network-level predictions in controlled intervention studies. To our knowledge, this work represents the first application of in silico network perturbation to human exercise biology, integrating genetically anchored habitual physical activity with graph-based intervention modelling and ageing clock evaluation.

## Results

### Causal multi-omic prioritisation of exercise-associated genes

We first leveraged genetically proxied habitual vigorous physical activity (VPA) to prioritise exercise-responsive molecular features using linkage disequilibrium (LD)-aware, overlap-aware MR across four orthogonal omic layers: plasma proteomics, whole blood DNA methylation, plasma glycomics, and blood single-cell transcriptomics. This analysis builds on our previously described supervised MR-Graph Attention Network (MR-GAT) framework for exercise gene prioritisation, in which graph neural networks were explicitly trained to predict MR-derived targets, but is summarised here to provide context for subsequent label-free *in silico* perturbation analyses (Juan et al., 2026, preprint).

Across molecular layers, MR signals were dense but unevenly distributed. In the proteomic layer, 2,304 out of 2,991 VPA single nucleotide polymorphism (SNP)-matched proteins (77.0%) demonstrated nominal evidence of causal association (p < 0.05). Similarly strong signal density was observed for epigenomic and glycomic traits, with 151 of 184 CpGs (82.1%) and 101 of 117 plasma glycan traits (86.3%) showing evidence of causal association with habitual VPA. In contrast, MR signals in the single-cell transcriptomic layer were more cell-type specific, with 119 of 364 gene–cell type tests (32.7%) meeting nominal significance, most prominently in B cells, dendritic cells, and CD8^+^ T cells. To permit unified network analysis, all MR signals were harmonised to gene symbols, yielding 2,370 unique genes with causal support from at least one molecular layer. Of these, only 21 genes showed convergent evidence across two layers and a single gene across three layers, indicating limited cross-layer redundancy and motivating a network-level rather than gene-centric interrogation.

For downstream perturbation analyses, these genes were embedded within a human multi-omic interaction network and represented using self-supervised contrastive graph representation learning, followed by MR-informed masked multi-task embedding refinement across proteomic, epigenomic, glycomic, and single-cell transcriptomic effect sizes. Importantly, no ageing-related targets or clock annotations were used during representation learning or refinement, ensuring that subsequent perturbation analyses were not biased toward ageing clock definitions.

### In silico perturbation reveals hierarchical network roles of exercise-responsive genes

We next performed targeted acute *in silico* perturbation of MR-prioritised genes by propagating single-event localised perturbations through the gene network using personalised PageRank diffusion. Because the underlying graph structure and embeddings were learned from MR-GAT modelling of genetically proxied habitual VPA, they encode time-averaged, long-term exercise adaptation; targeted *in silico* perturbation then simulates a single acute intervention applied to this adapted background, allowing separation of transient stress buffering from retained ageing-related molecular organisation, while ageing clocks index stable downstream molecular state. This diffusion-based perturbation framework is unsupervised with respect to ageing outcomes, relying solely on network topology and learned gene representations. For each perturbed gene, downstream diffusion neighbourhoods were quantified and evaluated for enrichment of genes defining established biological ageing clocks—Horvath, Hannum, PhenoAge, DunedinPACE, and Proteomic Clock— using degree-matched null models.

Perturbation of seven experimentally validated glycosylation enzymes produced diffusion neighbourhoods significantly enriched for ageing clock-associated genes across multiple epigenetic clocks (empirical p < 0.01). These effects were robust to weighting by differential methylation magnitude and showed disproportionate alignment with the proteomic ageing axis, indicating that glycosylation networks occupy positions closely coupled to stable ageing signatures.

In contrast, perturbation of DNA repair and stress response genes—including those robustly prioritised by supervised MR-GAT modelling and experimentally validated following acute exercise in humans—did not yield consistent enrichment for ageing clock architecture (p > 0.1 across clocks). Diffusion from these genes was broader and less clock-specific, consistent with upstream roles in buffering acute physiological stress rather than encoding long-term molecular state.

### Experimental validation of network-predicted exercise responses in humans

To anchor in silico perturbation findings to biological responses *in vivo*, we examined acute molecular adaptations to a single bout of vigorous exercise. Participants completed a high-intensity exercise session (≈80% VO_2_max), with paired pre- and post-exercise sampling enabling assessment of plasma N-glycosylation traits and targeted expression of canonical genome maintenance and DNA repair genes. Experimental procedures and analytical workflows have been reported previously (Juan et al., 2025, preprint). Notably, the glycomic platform quantified the same plasma N-glycan traits represented in the multi-omic network, enabling direct comparison between experimental responses and network-level predictions.

Acute exercise elicited a coordinated plasma glycomic response characterised by reduced agalactosylation (G0) alongside increased mono- and di-galactosylation (G1, G2), elevated total galactosylation (Gtotal), and increased total sialylation (Stotal) (all *p* < 0.05). In contrast, bisecting GlcNAc abundance (Btotal) did not change acutely post-exercise. These patterns indicate selective modulation of terminal glycan features rather than global remodelling of glycan architecture. Importantly, galactosylation is among the most robustly replicated glycomic correlates of biological ageing in humans, with higher IgG and plasma galactosylation consistently associated with younger biological age and reduced inflammatory tone. The observed exercise-induced shift toward increased galactosylation therefore aligns directionally with established glycan-based ageing signatures rather than reflecting nonspecific stress responses.

At the network level, these glycomic changes converged with pathways prioritised by Mendelian randomisation and supervised graph learning. Among the highest-ranked exercise-associated genes identified by the MR-GAT framework, multiple enzymes governing galactosylation and sialylation were consistently represented, including *B4GALT1, ST6GAL1, ST3GAL1, ST3GAL4, MGAT3, MGAT5*, and *MGAT5B*, supporting a causal link between habitual physical activity, regulated glycan remodelling, and ageing-relevant molecular organisation.

In parallel, expression of key genome maintenance and DNA repair genes prioritised by MR-GAT—*SIRT1, PARP1*, and *RAD51*—increased significantly following acute exercise (all *p* < 0.05). These genes form a coordinated repair axis supporting high-fidelity homologous recombination and damage resolution during physiological stress. Collectively, these experimental findings demonstrate concordance between network-prioritised pathways and acute human exercise responses at the level of pathway engagement rather than uniform gene-level effects. Acute exercise preferentially modulated terminal glycosylation features while activating DNA repair programmes, providing biological context for the distinct network roles revealed by subsequent in silico perturbation analyses.

### Network hierarchy links acute exercise responses to long-term ageing architecture

To characterise the network roles of exercise-responsive genes, we quantified the topological consequences of targeted *in silico* perturbation using complementary diffusion metrics. Diffusion concentration, measured by the Gini coefficient, captured the degree of localisation of perturbation effects, whereas diffusion entropy quantified regulatory breadth and network-wide signal dispersion (Figure 1).

**Figure 1.**
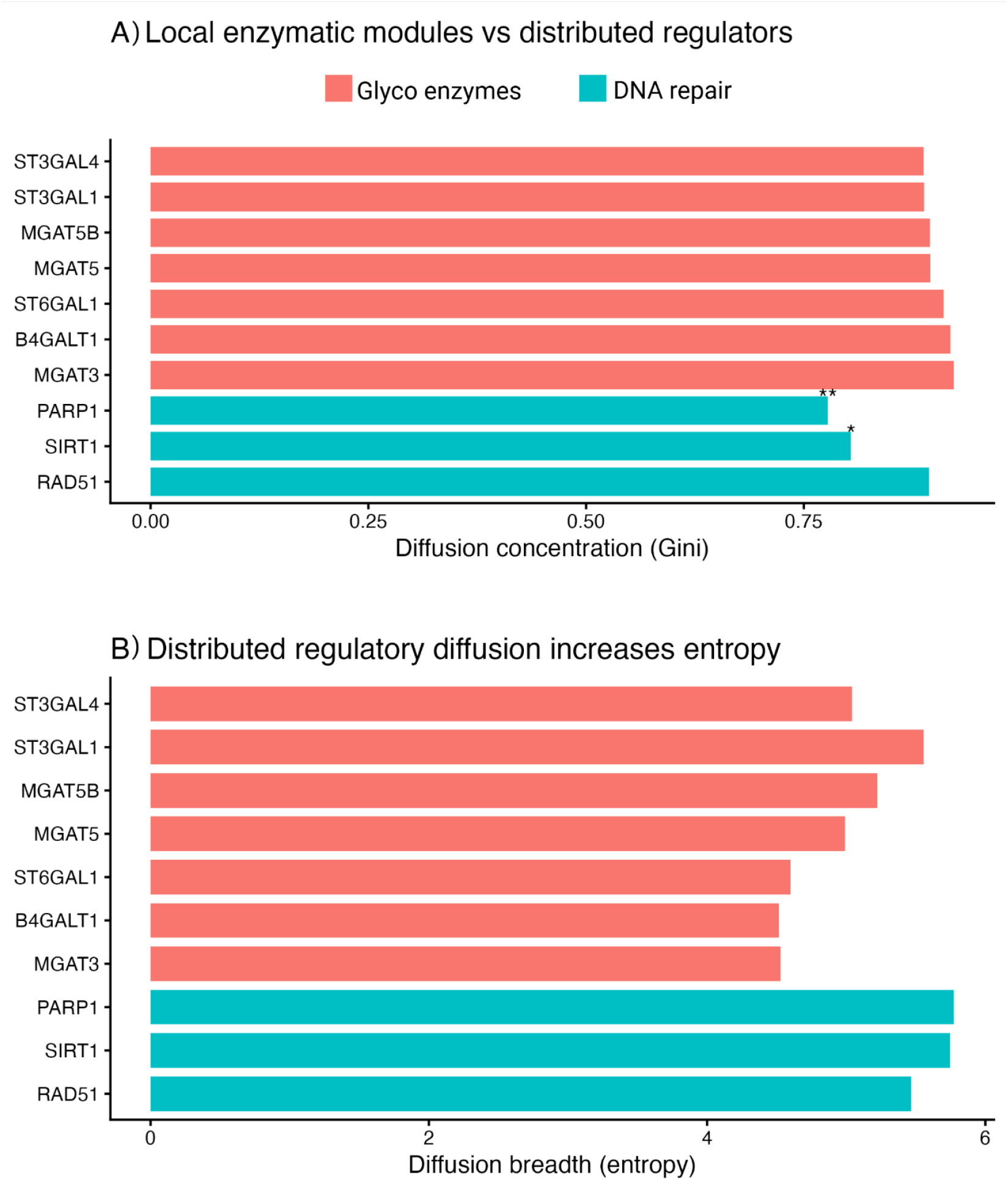
Distinct diffusion signatures distinguish local enzymatic modules from distributed regulatory genes. A) Diffusion concentration (Gini coefficient) following targeted acute in silico perturbation of exercise-responsive genes. Glycosylation enzymes exhibit highly concentrated diffusion profiles, consistent with localised, modular network influence, whereas DNA repair and regulatory genes display lower concentration, indicating broader signal propagation. B) Diffusion breadth (entropy) is increased following perturbation of DNA repair and regulatory genes compared with glycosylation enzymes, reflecting more distributed network engagement. Statistical annotations indicate differences relative to glycosylation enzymes (*p < 0.05, **p < 0.01; empirical permutation tests). This pattern indicates that glycosylation enzymes operate as local enzymatic modules, exerting focused influence over tightly connected network neighbourhoods. Notably, these neighbourhoods were significantly enriched for genes defining multiple epigenetic ageing clocks and showed disproportionate alignment with the proteomic ageing axis, consistent with a role in encoding stable molecular state.

Perturbation of validated glycosylation enzymes consistently produced highly localised diffusion profiles, reflected by elevated Gini coefficients and comparatively lower entropy (Figure 1A–B). These patterns indicate that glycosylation enzymes operate as compact enzymatic modules, exerting focused influence over tightly connected network neighbourhoods. Notably, these neighbourhoods were significantly enriched for genes defining multiple epigenetic ageing clocks and showed disproportionate alignment with the proteomic ageing axis, consistent with a role in encoding stable, system-level molecular state.

In contrast, perturbation of DNA repair and stress-response genes—including *PARP1, SIRT1*, and *RAD51*—produced broad, low-concentration diffusion patterns characterised by increased entropy (Figure 1B). Although these genes occupied central positions within the network, their influence was distributed across diverse functional modules rather than concentrated within ageing clock-enriched regions. This topology is consistent with upstream regulatory roles in coordinating acute exercise-induced stress responses rather than specifying long-term molecular state. Network visualisation of diffusion neighbourhoods reinforced this distinction (Figures 2 and 3). Glycosylation enzymes were embedded within compact subnetworks densely populated by ageing clock-associated genes (Figure 2), whereas DNA repair genes resided within expansive, highly interconnected regions spanning transcriptional regulation, signalling, chromatin organisation, and stress-response pathways (Figure 3).

**Figure 2.**
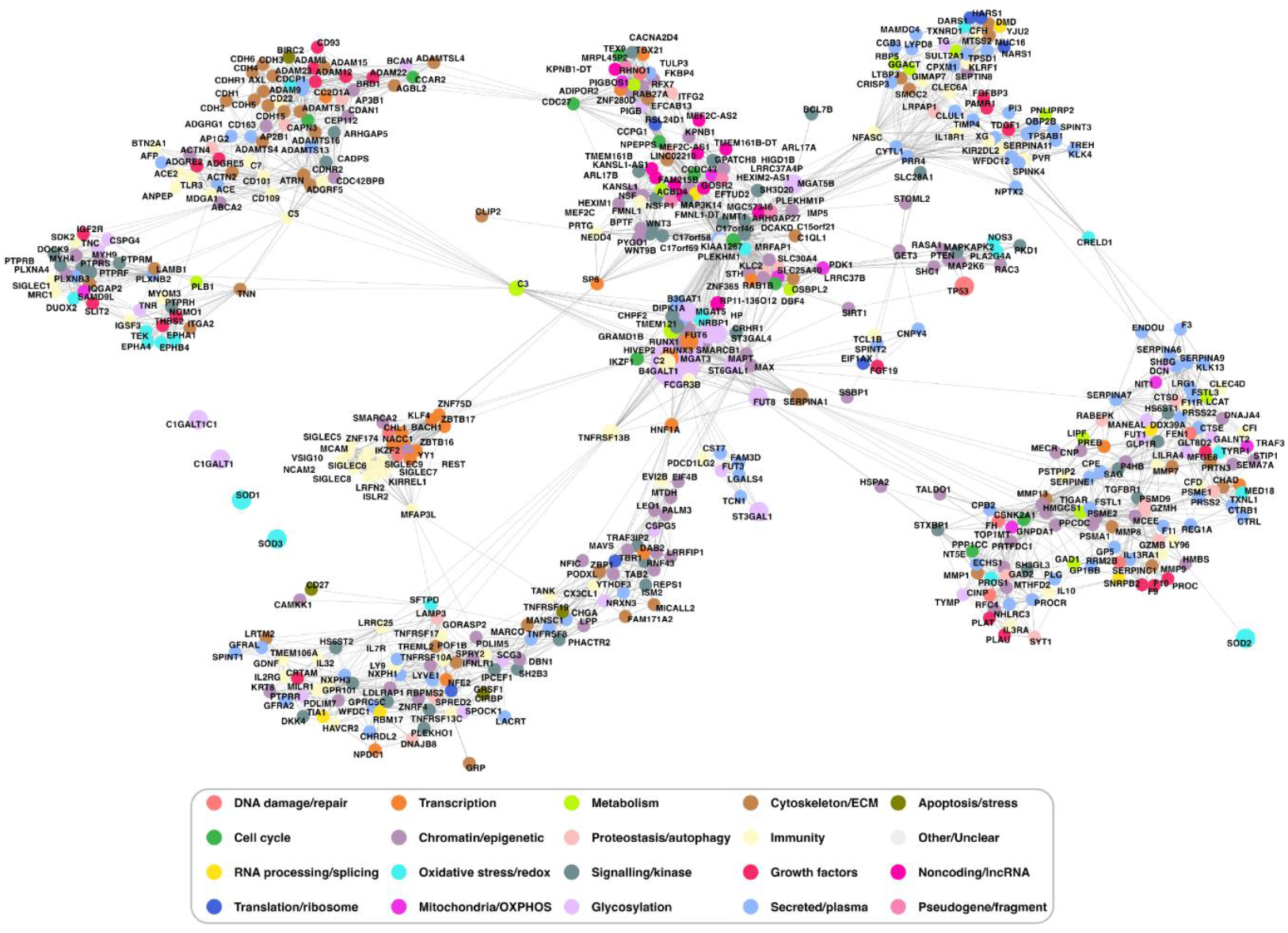
Glycosylation enzymes form locally concentrated network modules aligned with ageing clock architecture. The top 500 genes with ageing clock support are shown in their network neighbourhood centred on the glycosylation enzyme B4GALT1, illustrating the topological organisation of glycosylation-related genes within the exercise-responsive molecular network. Nodes represent genes coloured by functional category, and edges indicate network connectivity. Glycosylation enzymes cluster within compact, highly interconnected local modules, consistent with concentrated diffusion and preferential enrichment of ageing clock-associated genes following perturbation. This localised architecture highlights glycosylation pathways as proximal encoders of stable molecular states captured by biological ageing clocks.

**Figure 3.**
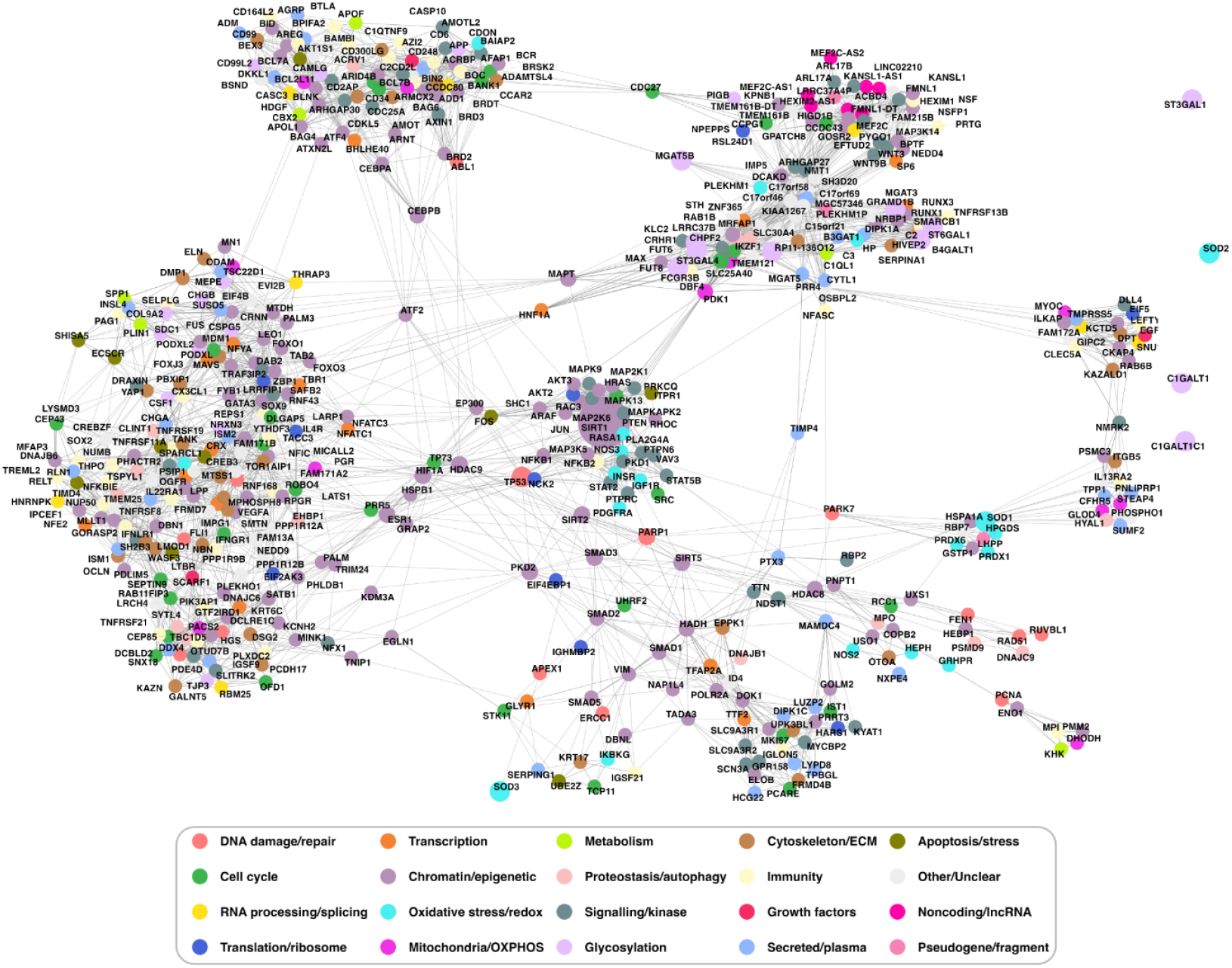
DNA repair and stress response genes occupy distributed regulatory positions within the exercise-responsive network. The top 500 genes with ageing clock support are shown in their network neighbourhood centred on SIRT1 and DNA repair genes (PARP1, RAD51). Nodes represent genes coloured by functional category, and edges denote network connectivity. In contrast to the compact topology observed for glycosylation enzymes, DNA repair and stress response genes are embedded within distributed regulatory regions. This diffuse topology is consistent with widespread diffusion, high entropy, and limited enrichment for ageing clock genes, reflecting upstream roles in buffering acute physiological stress rather than encoding long-term ageing-related molecular states.

**Figure 4.**
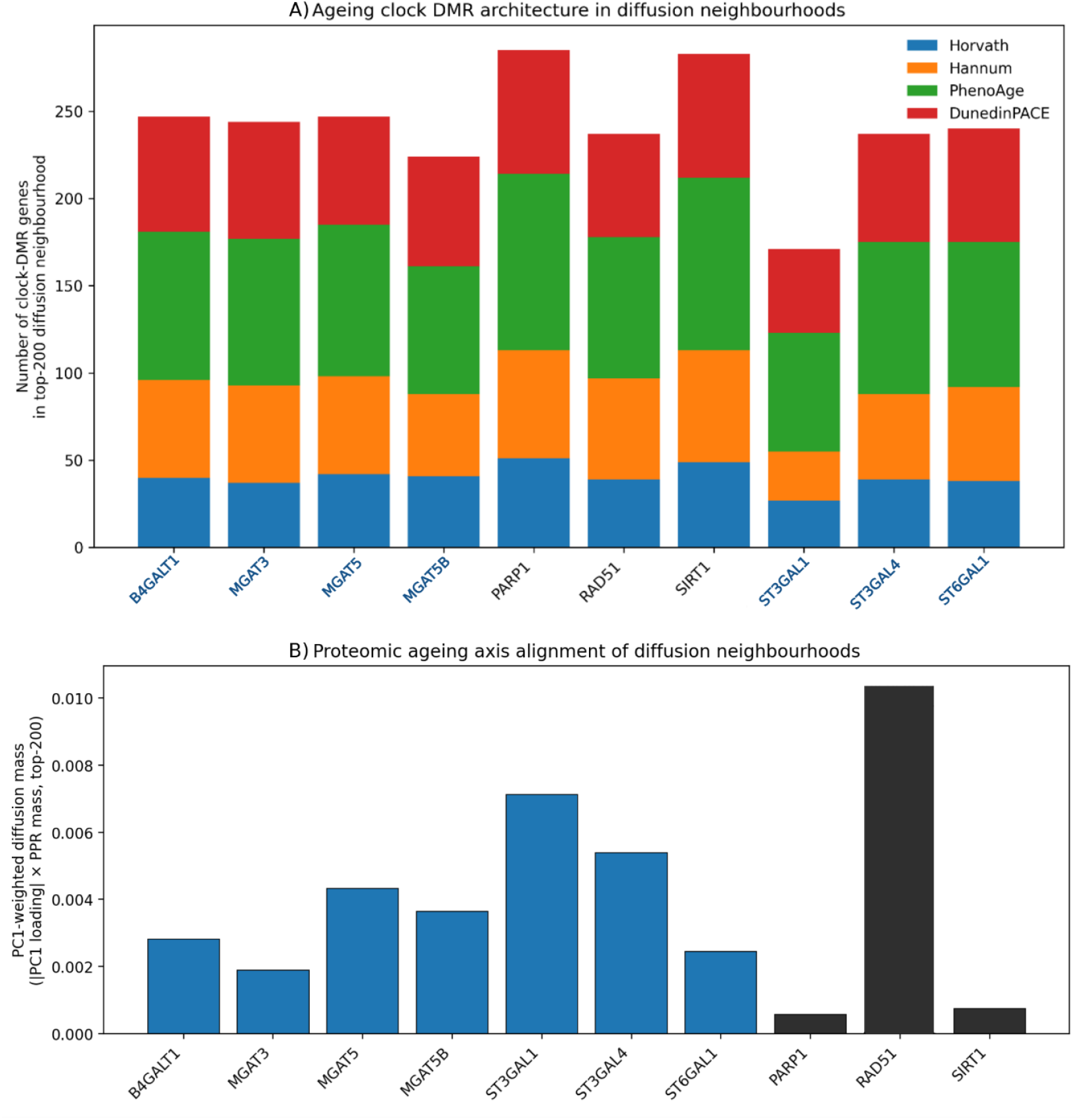
Diffusion-based impact of exercise-associated genes on epigenetic and proteomic ageing architecture distinguishes diffusion breadth from ageing-state alignment. A) Stacked bar plot showing the total number of epigenetic ageing clock DMR-associated genes captured within the top-200 diffusion neighbourhood of each perturbed gene, stratified by clock (Horvath, Hannum, PhenoAge, DunedinPACE). Gene labels are colour-coded by functional class: glycosylation enzymes (blue) and DNA repair/stress-response genes (black). DNA repair genes exhibit broader diffusion neighbourhoods with higher overall DMR coverage across clocks, reflecting their central, upstream roles in coordinating acute stress and repair responses. In contrast, glycosylation enzymes capture fewer but more clock-structured subsets of DMR targets, indicating more selective engagement with ageing-related molecular architecture. B) Bar plot showing alignment with a proteomic ageing axis, quantified as diffusion mass within the top-200 neighbourhood weighted by absolute consensus PC1 loadings. Despite lower total DMR coverage in panel A, glycosylation enzymes show consistently elevated PC1-weighted diffusion signals across multiple genes, indicating coherent coupling to stable proteomic ageing structure. DNA repair/stress-response genes display heterogeneous and gene-specific alignment, including a single dominant peak (RAD51), consistent with broad but non-specific network influence. Together, these panels demonstrate that DNA repair pathways exert widespread diffusion (breadth) without selectively encoding ageing state, whereas glycosylation networks exhibit more focused diffusion that preferentially aligns with epigenetic and proteomic ageing clocks, supporting their role as downstream integrators of stable molecular ageing architecture.

Together, these analyses reveal a hierarchical organisation within exercise-responsive molecular networks. DNA repair pathways act as distributed regulators that preserve genome integrity during acute exercise-induced stress, whereas glycosylation networks function as localised effectors that encode the stable molecular patterns captured by biological ageing clocks. This hierarchy reconciles experimental evidence for exercise-induced DNA repair activation with the relative absence of DNA repair genes from ageing clock definitions, providing a mechanistic bridge linking habitual physical activity to long-term biological ageing trajectories without conflating acute repair responses with molecular ageing itself.

### Impact of in silico perturbation on epigenetic and proteomic ageing clock architecture

To quantify how exercise-associated genes influence molecular ageing architecture, we assessed the downstream impact of *in silico* perturbation on genes defining established epigenetic and proteomic ageing clocks. For each perturbed gene, diffusion neighbourhoods generated by personalised PageRank were evaluated for enrichment of clock-associated differentially methylated region (DMR) genes across four epigenetic clocks (Horvath, Hannum, PhenoAge, DunedinPACE), as well as alignment with a proteomic ageing axis defined by first principal component (PC1) loadings of a Proteomic Clock derived from various cohorts. Significance was assessed using degree-matched null models with empirical permutation testing, ensuring that observed effects were not attributable to network connectivity alone.

Perturbation of validated glycosylation enzymes produced a consistent and significant impact on ageing clock architecture. Diffusion neighbourhoods originating from glycosylation genes captured a disproportionately large fraction of epigenetic clock DMR genes across multiple clocks, exceeding degree-matched expectations (empirical *p* < 0.01). These effects were robust across clocks that differ in biological emphasis, including mitotic history (Horvath), immune ageing (Hannum), and morbidity-linked phenotypes (PhenoAge, DunedinPACE). In parallel, glycosylation gene perturbations showed strong alignment with the proteomic ageing axis, reflected by elevated diffusion mass weighted by absolute PC1 loadings, indicating close coupling between glycosylation network structure and stable proteomic ageing signatures.

In contrast, perturbation of DNA repair and stress response genes—including *PARP1, SIRT1*, and *RAD51*—did not yield consistent enrichment of epigenetic clock DMR genes or significant alignment with the proteomic ageing axis (empirical *p* > 0.1 across clocks). Although these genes were centrally positioned within the exercise-responsive network and exhibited broad diffusion profiles, their downstream influence was distributed across diverse functional modules rather than concentrated within clock-associated regions. This pattern indicates that DNA repair pathways, while essential for coordinating acute stress responses, do not directly organise the stable molecular features captured by ageing clocks. Together, these findings demonstrate that *in silico* perturbation distinguishes exercise-associated genes that predictively shape ageing clock architecture from those that primarily mediate transient physiological responses. Glycosylation networks emerge as proximal organisers of long-term molecular ageing states, whereas DNA repair pathways function as upstream regulators that preserve genomic integrity without directly encoding ageing clock structure.

## Discussion

Exercise modulates biological ageing, yet the molecular hierarchy linking habitual physical activity to long-term ageing signatures in humans remains unclear. Addressing this gap requires moving beyond causal gene identification toward frameworks that can predict how exercise-responsive pathways reorganise molecular systems over time. In this study, we extend genetically anchored causal prioritisation with predictive, network-based *in silico* perturbation to forecast how exercise-associated genes propagate influence through molecular networks to shape ageing-relevant molecular architecture. By integrating MR, self-supervised graph representation learning, diffusion-based perturbation, and biological validation in a controlled human exercise intervention, we provide a unified framework that links acute exercise responses to stable ageing clock organisation without conflating transient stress signalling with biological ageing itself. Importantly, modelling acute perturbations within a network shaped by habitual exercise is a biologically appropriate abstraction, as ageing clocks capture retained molecular organisation rather than short-lived stress responses.

Consistent with prior work, genetically proxied habitual VPA was associated with widespread molecular effects across proteomic, epigenomic, glycomic, and single-cell transcriptomic layers. However, harmonisation of these signals to gene symbols revealed limited cross-layer convergence, with only a small subset of genes supported by multiple omic layers. This sparse overlap highlights a key limitation of gene-centric interpretations of exercise MR and is consistent with observational transcriptomic and multi-omic analyses of exercise responses, which report physiological adaptation accompanied by extensive molecular heterogeneity and limited gene-level replication across tissues, time points, and omic layers (MoTrPAC Study Group, 2024; Voisin et al., 2024). Together, these observations motivate a network-based interrogation in which exercise-responsive genes are interpreted in terms of their position, connectivity, and downstream influence, rather than as isolated effect sizes.

Embedding MR-prioritised genes within a human multi-omic interaction network was performed using self-supervised contrastive graph representation learning, followed by masked multi-task embedding refinement using MR effect sizes across proteomic, epigenomic, glycomic, and single-cell transcriptomic layers, without incorporating any ageing-related label. This design ensured that subsequent associations with ageing clocks emerged from network topology and information flow rather than supervised optimisation toward ageing outcomes. Targeted *in silico* perturbation then provided a predictive lens through which to examine how exercise-responsive genes propagate influence across molecular systems. This approach builds on established principles from network biology demonstrating that disease and trait-relevant molecular effects localise to network modules rather than single genes (Goh et al., 2007; Menche et al., 2015), and that network influence is not equivalent to node degree or centrality (Kitsak et al., 2010).

Across multiple epigenetic ageing clocks and a proteomic ageing axis, perturbation of validated glycosylation enzymes consistently produced diffusion neighbourhoods enriched for clock-defining genes, exceeding degree-matched expectations and showing strong alignment with proteomic ageing loadings. In contrast, perturbation of canonical DNA repair and stress response genes—including *PARP1, SIRT1*, and *RAD51*—did not yield consistent enrichment of ageing clock architecture, despite their central network positions and broad diffusion profiles. Topological analyses clarified the basis of this divergence. Glycosylation enzymes occupied compact, highly localised network modules characterised by high diffusion concentration and low entropy, indicating focused influence over tightly connected neighbourhoods. DNA repair genes, by contrast, resided within distributed regulatory regions spanning transcriptional control, chromatin organisation, metabolic signalling, and stress-response pathways, consistent with broad but non-specific downstream propagation. These results establish a hierarchical network organisation in which glycosylation pathways act as proximal encoders of ageing-related molecular state, while DNA repair pathways function as upstream buffers of acute physiological stress.

This hierarchy is consistent with theoretical models that frame ageing as an emergent consequence of selection-driven homeostatic reorganisation rather than cumulative molecular damage. In the four-process model proposed by Wordsworth et al. (2026), DNA damage responses and genome maintenance pathways are conceptualised as adaptive mechanisms repeatedly activated by physiological stress to preserve tissue viability and suppress runaway selection (*i*.*e*. the progressive amplification of cell-intrinsic growth or survival advantages that, if left unchecked, lead to clonal expansion, tissue dysfunction, and loss of organism-level homeostasis) rather than as direct drivers of ageing. This framework builds on and extends earlier critiques of damage-centric ageing theories and aligns with hyperfunction and quasi-program models that emphasise dysregulated growth and homeostasis over molecular damage accumulation (Gladyshev, 2016; Blagosklonny, 2013). A central motivation for Wordsworth’s view is the paradox revealed by hormesis, whereby molecular damage can increase while lifespan improves, demonstrating that damage responses cannot themselves encode ageing states. Ageing is instead proposed to emerge downstream of stress buffering, through slower reorganisation of metabolic homeostasis, with secondary immune and inflammatory consequences that give rise to stable molecular states

Recent causality-enriched epigenetic ageing models provide independent support for this distinction. Ying et al. (2024) introduced DamAge and AdaptAge, two DNA methylation-based ageing clocks constructed from CpG loci whose methylation levels exert deleterious or protective causal effects on ageing-related outcomes, respectively, as inferred by epigenome-wide MR. These clocks are derived exclusively from CpG methylation measurements and therefore capture persistent regulatory epigenetic states rather than DNA damage itself or the activity of genome maintenance and repair pathways. Importantly, the causal separation of damaging (DamAge) and adaptive (AdaptAge) epigenetic signals resolves why DNA damage response pathways—despite their central role in stress resilience—show limited alignment with ageing-clock DMRs: adaptive repair responses are repeatedly engaged and successful repair leaves little retained molecular imprint, whereas DamAge CpGs capture persistent downstream consequences of damage exposure that are not fully resolved by adaptation. This distinction explains why DNA repair genes exert broad network influence in our perturbation analyses yet do not define ageing-clock architecture, whereas integrative pathways such as glycosylation align more closely with retained ageing signatures.

The controlled human high-intensity exercise intervention provides critical biological context for this hierarchy. Acute exercise elicited rapid modulation of terminal plasma N-glycosylation features, including reduced agalactosylation and increased galactosylation and sialylation, while core architectural features such as bisecting GlcNAc remained unchanged, consistent with dynamic regulation of immune and inflammatory signalling rather than wholesale glycan remodelling. In parallel, expression of genome maintenance and DNA repair genes prioritised by the network—including *SIRT1, PARP1*, and *RAD51*—increased acutely following high-intensity exercise, confirming engagement of a coordinated repair programme *in vivo*. At the pathway level, experimental findings converged with network predictions: enzymes governing galactosylation and sialylation were both acutely responsive and centrally positioned within ageing-aligned network modules, whereas enzymes controlling glycan architecture showed no acute change, consistent with their inferred role in shaping longer-term glycosylation capacity.

Together, these results clarify how acute exercise-induced stress is translated into durable molecular resilience without being directly embedded within biological ageing clocks. DNA repair pathways act as essential, adaptive buffers that preserve genome integrity across repeated physiological challenges but remain diffuse and non-specific at the level captured by ageing clocks. In contrast, glycosylation networks occupy a downstream, integrative position, encoding immune-metabolic consequences of habitual physical activity that accumulate across repeated exposures and are consequently represented within epigenetic and proteomic ageing measures. By introducing graph-based *in silico* perturbation to human exercise biology, this study demonstrates how predictive network modelling—akin to controllability and influence frameworks in complex systems biology (Liu et al., 2011; Kitsak et al., 2010)—can complement causal inference and experimental validation. Rather than producing static gene lists, this approach enables forecasting of emergent molecular states, offering a principled route to identify pathways through which lifestyle exposures shape long-term ageing trajectories.

Beyond its biological implications, this study contributes a generalisable methodological framework for causal systems inference in human exercise biology. Prior exercise genomics studies have generated rich single and multi-omic atlases of training-induced molecular change across tissues and time, typically relying on differential analysis, correlation with phenotypes or ageing clocks, and *post-hoc* pathway enrichment to interpret exercise responses. While these approaches have been instrumental in mapping exercise-responsive molecular landscapes, they remain largely descriptive and do not resolve how genetically anchored exercise signals are organised within interacting molecular networks, nor how local perturbations propagate to influence ageing-relevant molecular architecture. In contrast, our framework integrates causally informed multi-omic MR with self-supervised graph representation learning, followed by masked multi-task embedding refinement across proteomic, epigenomic, glycomic, and single-cell transcriptomic effect sizes, and diffusion-based *in silico* perturbation. This design enables prediction of how causally prioritised exercise-responsive pathways reorganise molecular networks, rather than merely cataloguing correlated genes or enriched pathways. By performing acute perturbation within a network shaped by genetically proxied habitual VPA, the approach bridges causal inference and predictive systems biology, providing a principled means to interrogate how lifestyle exposures shape ageing-relevant molecular organisation.

Several future directions follow naturally from this work. While perturbation analyses were intentionally restricted to single-event (acute) interventions, extension to repeated or temporally weighted perturbation schemes may enable closer approximation of cumulative exposure as longitudinal multi-omic datasets become available. Incorporation of tissue-specific or cell-type-resolved interaction networks will further refine context-dependent ageing mechanisms, particularly beyond blood-derived omics. In addition, integration of other genetically proxied exposures—such as dietary factors, pharmacological interventions, or disease liabilities—would allow comparative evaluation of how distinct interventions reshape ageing-related molecular architecture within a unified network scaffold. Finally, prospective validation in longitudinal exercise or lifestyle intervention cohorts will be essential to test whether pathways predicted to align with ageing clocks also predict long-term functional and clinical outcomes.

Our perturbation-based network analyses do not support a model in which DNA damage responses directly encode biological ageing state. Instead, viewed through the lens of the DNA damage theory of ageing, these findings indicate that genome instability and repair capacity are necessary determinants of physiological stress tolerance, but are insufficient to specify ageing state itself. Ageing-relevant molecular architecture appears to emerge downstream of repeated stress buffering, through slower, integrative regulatory processes that stabilise system-level metabolic and immune organisation over time. Consistent with this interpretation, genome maintenance pathways act as adaptive buffers of physiological stress, whereas ageing-relevant molecular states are captured by time-integrated regulatory programmes rather than by acute damage responses.

## Methods

### Graph structure and node representations

All *in silico* perturbation analyses were performed on the same gene interaction graph and node index used in the supervised MR-GAT model, ensuring full topological consistency with the primary exercise-ageing network (Juan & Ntasis, 2026). Nodes represent protein-coding genes, and edges encode functional gene-gene associations derived from the k-nearest neighbors (KNN) similarity graph constructed from integrated multi-omic, structural, and functional features, as described previously. The graph topology was treated as fixed throughout all perturbation and diffusion analyses. Each gene node was associated with a learned low-dimensional embedding vector. Base embeddings were obtained via self-supervised contrastive pretraining, followed by MR-informed masked multi-task embedding refinement. These embeddings provide a continuous latent representation of gene context, integrating functional annotation, structural properties, and network neighbourhood information.

### Self-supervised contrastive embedding pretraining

To obtain task-agnostic gene embeddings independent of explicit ageing or MR supervision, we performed self-supervised contrastive pretraining on the node feature matrix. Two stochastic views of each node were generated via feature-level dropout and encoded using a shared multilayer perceptron (MLP). Embeddings were trained using a SimCLR-style NT-Xent contrastive loss, which encourages embeddings of the same gene under stochastic feature dropout to be similar while maximising separation from all other genes in the batch. Training was performed for 500 epochs using the Adam optimiser (learning rate = 1×10^−3^, weight decay = 1×10^−5^). The resulting embeddings capture intrinsic graph-aware functional similarity without reference to ageing clocks or causal targets. Full loss definition and optimisation details are provided in the supplementary files.

### MR-informed embedding refinement via masked multi-task regression

To align the latent embedding space with causal exercise biology while preserving global graph structure, embeddings were refined using masked multi-task regression on standardised MR effect sizes across all omic layers simultaneously, with one task-specific regression head per omic layer. A lightweight projection network mapped base embeddings ***Z*** to refined embeddings ***Z′***, which were then used to predict MR effect sizes for each omic layer via the corresponding regression heads.

MR targets were sparse and unevenly distributed across layers; therefore, missing targets were masked such that loss was computed only for genes with valid MR estimates in the corresponding layer. A Huber loss was used for robustness to outliers, and a stability regularisation term penalised large deviations between ***Z*** and ***Z′***, preserving the topology learned during self-supervised pretraining.

This formulation enables embeddings to encode shared and coordinated causal signals across omic layers, allowing genes with consistent multi-layer effects to emerge as cross-omic integrators while retaining sensitivity to layer-specific regulation. All training objectives, masking logic, and optimisation details are provided in the supplementary files.

### In silico gene perturbation in embedding space

*In silico* perturbation was implemented by explicitly modifying the embedding vector of a target gene while holding all other embeddings fixed. For a given gene ***g*** with embedding **z**_***g***_, perturbation was defined as:

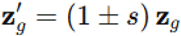

where ***s*** is the perturbation strength (default ***s*** = 1.0), with + corresponding to overexpression (OE) and − to knockdown (KD). The magnitude of the perturbation was quantified as:

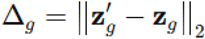

This scalar ensured that downstream network effects scaled with perturbation size rather than direction alone. The embedding-space perturbation framework enables controlled *in silico* simulation of gene-level interventions, analogous to knockdown or overexpression experiments, without altering graph topology or retraining the network. All perturbation analyses model single-event (acute) gene-level interventions; cumulative, repeated, or time-resolved perturbation dynamics were not simulated.

### Interpretation of acute perturbation on a habitual exercise-adapted embedding space

The learned gene embeddings and network topology are derived from MR of genetically proxied habitual VPA, and therefore represent a time-averaged, long-term molecular adaptation to repeated exercise exposure rather than an instantaneous physiological state. Acute *in silico* perturbation is applied to this adapted background to simulate a single local intervention (e.g., transient gene activation or suppression) occurring within an already exercise-conditioned system. Mathematically, this corresponds to perturbing a steady-state latent representation rather than re-estimating the embedding space itself. Network diffusion then quantifies whether such local deviations propagate preferentially toward regions encoding stable ageing-related molecular architecture, as indexed by epigenetic and proteomic ageing clocks. This formulation enables separation of transient stress buffering from retained system-level organisation, without conflating acute molecular responses with the embedding of long-term biological ageing state.

### Network diffusion of perturbation effects

To propagate local perturbations across the gene interaction network, we applied a personalised PageRank (PPR)-style diffusion process centred on the perturbed gene. Starting from a one-hot seed vector **e**_***g***_, diffusion was iteratively updated as:

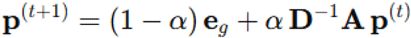

where α is the restart probability (default α = 0.85), **A** is the adjacency matrix, and **D** is the diagonal degree matrix. Diffusion was run for 50 iterations, yielding a steady-state influence vector **p** over all genes. The final diffusion-weighted impact score for each gene *i* was defined as:

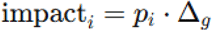

Genes were ranked by this score to identify those most affected by the simulated perturbation. This diffusion process yields a static steady-state influence profile capturing first-order network propagation of a single perturbation event.

### Quantifying perturbation breadth and concentration

To characterise whether perturbations produced localised or global network effects, we quantified the breadth of diffusion using complementary metrics. Diffusion entropy was computed as:

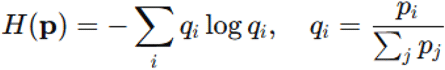

where higher entropy indicates broader network propagation. In parallel, the Gini coefficient of **p** was computed to quantify inequality of diffusion mass across nodes. Top-K mass concentration (K = 10, 50, 200) was also recorded.

### Degree-matched null models and empirical significance testing

To assess whether observed diffusion patterns exceeded expectations given network topology, degree-matched null distributions were constructed. For each perturbed gene, null seed genes with similar degree were sampled, and diffusion was recomputed using identical parameters. Two-sided empirical p-values were computed for all diffusion metrics as:

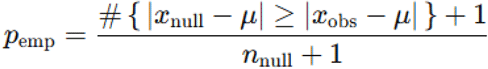

where *x*_obs_ is the observed statistic, *x*_null_ are null values, and μ is the null mean. This non-parametric framework controls for degree-driven and topological confounding.

### Quantifying impact on ageing-related molecular programmes

To determine whether simulated interventions preferentially reorganised ageing-related molecular architecture rather than generic network neighbourhoods, diffusion footprints were intersected with curated ageing-associated gene sets. Diffusion footprints were further intersected with ageing-related gene sets, including epigenetic clock DMR genes (Horvath, Hannum, PhenoAge, DunedinPACE), proteomic ageing PC1 genes, and a validated glycosylation enzyme panel. For each annotation set ***S***, total and top-K captured diffusion mass were computed as:

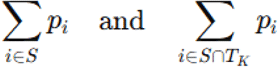

Optionally, DMR-based analyses incorporated per-gene weights derived from the number of DMRs or mean absolute methylation change. All annotation-level metrics were evaluated against degree-matched null distributions using the empirical testing framework above.

### External evaluation using independent ageing targets

To independently validate whether learned gene embeddings encode ageing-relevant biological structure, external ageing prediction tasks were performed using frozen embeddings. Importantly, no ageing-related labels were used during self-supervised pretraining, MR-informed refinement, or perturbation analysis, ensuring strict separation between representation learning and evaluation.

Embeddings were evaluated against independent ageing-related targets, including epigenetic clock DMR summaries and proteomic ageing principal components (PC1). Regularised linear models (Ridge or Elastic Net) were trained on predefined degree-stratified splits to control for network centrality. Performance was assessed using held-out test-set *R*^*2*^, Spearman rank correlation, and permutation-based significance testing. This external evaluation tests whether the learned embedding space captures ageing-relevant molecular organisation beyond causal exercise supervision alone.

## Data Availability

All data produced are available online at https://github.com/cgjuan01

## Data Availability

Supplementary files are available at https://github.com/cgjuan01

## Code Availability

All scripts used for data processing, MR analyses, network construction, and GNN training are available at https://github.com/cgjuan01

